# The minor allele of the CREBRF rs373863828 p.R457Q coding variant is associated with reduced levels of myostatin in males: Implications for body composition

**DOI:** 10.1101/2021.07.13.21260462

**Authors:** Kate Lee, Sanaz Vakili, Hannah J. Burden, Shannon Adams, Greg C. Smith, Braydon Kulatea, Morag Wright-McNaughton, Danielle Sword, Conor Watene-O’Sullivan, Robert D. Atiola, Ryan G. Paul, Lindsay D. Plank, Phillip Wilcox, Prasanna Kallingappa, Tony R. Merriman, Jeremy D. Krebs, Rosemary M. Hall, Rinki Murphy, Troy L. Merry, Peter R. Shepherd

## Abstract

The minor allele (A) of the rs373863828 variant (p.Arg457Gln) in CREBRF is restricted to indigenous peoples of the Pacific islands (including New Zealand Māori and peoples of Polynesia), with a frequency up to 25% in these populations. This allele associates with a large increase in body mass index (BMI) but with significantly lower risk of type-2 diabetes (T2D). It is unclear whether the increased BMI is driven by increased adiposity or by increased lean mass. Hence, we undertook body composition analysis using DXA in 189 young men of Māori and Pacific descent living in Aotearoa New Zealand. The rs373863828 A allele was associated with a trend toward increased relative lean mass although this was not statistically significant (p=0.06). Notably though this allele was associated with significantly lower circulating levels of the muscle inhibitory hormone myostatin (p<0.05). This was further investigated in two Arg458Gln knockin mouse models on FVB/Nj and C57Bl/6j backgrounds. Supporting the human data, significant increases in relative lean mass were observed in male knockin mice. This was more significant in older mice (p<0.01) where it was associated with increased grip strength (p<0.01) and lower levels of myostatin (p <0.05). Overall these results provide new evidence that the rs373863828 A-allele is associated with a reduction of myostatin levels which likely contributes to increased lean muscle mass component of BMI, at least in males.

## INTRODUCTION

A large number of genetic variants have been identified that contribute to the risk of developing either type-2 diabetes (T2D) [1–3] or obesity [3–5]. However, the vast majority of common gene variants only have a small effect size. One exception is the rs373863828 variant in the *CREBRF* gene. This gene, which codes for the highly conserved, but poorly understood, protein CREBRF (CREB3 Regulatory Factor, also known as Luman Recruitment Factor (LRF) of C5orf41) [6]. The rs373863828 minor (A) allele causes the p.Arg457Gln substitution and the functional consequences of this are not well understood. This variant is unique to isolated populations in the islands of the Pacific where it is present at minor allele frequencies (MAF) of up to 25% in Polynesia (including Samoa, Aotearoa NZ Māori) [7–9] and at lower MAF in some areas of Melanesia and Micronesia [9, 10]. Importantly, of single gene variants common in a population it has the biggest impact on BMI, being associated with an increase of up to 3 kg/m^2^ per rs373863828 A allele [7–10]

Increases in BMI in adults are commonly associated with a relative increase in adiposity [11] and initial evidence suggested the impact of the rs373863828 A allele on BMI was due to increased adiposity [8]. Based on this it was suggested this variant was an evolutionary adaptation for facilitating energy storage and thus was acting as a thrifty gene [8]. However, this would not be consistent with Māori and other peoples from Polynesia having higher relative lean mass for a given BMI compared to people of European or Asian ancestry [12, 13]. Further, recent follow-on studies undertaken in Samoa have indicated that in males the rs373863828 A allele is associated with increased lean mass, not adiposity, suggesting the effects on BMI may be driven by greater increases in lean mass rather than fat mass [14, 15]. To address these inconsistencies we examined the impact of the minor allele of rs373863828 p.Arg457Gln CREBRF on body composition in a cohort of young Māori and Pacific men and in two mouse knockin models.

## MATERIALS AND METHODS

### Human Studies

Healthy men of NZ Māori and / or Pacific Island descent (n=189) were recruited in Auckland and Wellington, Aotearoa New Zealand and gave written informed consent to participate. Eligibility criteria included being free from chronic illnesses including cardiovascular or metabolic disease, aged 18-45 years with a BMI of 20-45 kg/m^2^. This study was approved by the Health and Disability Ethics Committee, New Zealand (17STH79).

Body composition was determined by dual energy x-ray absorptiometry (DXA, model iDXA, GE Healthcare, Madison, WI and Hologic Horizon A, Hologic, Marlborough, MA). Resting energy expenditure (REE) was measured in overnight fasted participants by indirect calorimetry (Parvo Medics True One 2400, UT, USA and Promethion, Sable Systems, NV, USA) using a ventilated hood following 30 minutes resting in a supine position. An overnight fasted venous blood sample was collected between 8:00 am and 10 am in the morning with plasma collected in a tube containing EDTA. Plasma and serum were recovered before being frozen at −80°C. Circulating myostatin/GDF-8 in both human (plasma) and mouse (serum) samples was measured by commercial ELISA (R&D systems, Minneapolis, MN, USA, catalogue number RDSDGDF80). DNA was from whole blood isolated using a commercially available kit (DNA extraction minikit, Qiagen, Hilden Germany). *rs373863828* genotype was determined using a custom-designed Taqman probe-set (Applied Biosystems, Foster City, CA, USA) as described previously [16].

A multivariable linear regression model was used to test for associations with rs373863828 A allele (assuming an additive effect of the A-allele) with all parameters presented in Tables 2 (RStudio v1.2.1335 statistical software, www.rstudio.com). Proportion of self-reported grandparents of Polynesian ancestry (ANC), age and BMI were included as covariates where indicated [16].

### Mouse model

Orthologous mouse CREBRF p.458Q gene variant knock-in mice were generated on both FVB/NJ and C57Bl/6j backgrounds. The FVB mice were generated as previously reported [17]. The mice on the C57Bl/6j background were generated by the Rodent Genetics for Research (ROGER) Facility (Faculty of Medical and Health Sciences, University of Auckland, New Zealand) using CRISPR/Cas9 technology using the targeting strategy shown in Suppl Figure 1. Sequencing was used to verify the presence of the variant and lack of non-intended changes (sequencing primers AAGAACCCCATACCAGATAGC and TACAGTGACAAAGACAGGTATGG). Crispon was used to check the specificity of guide RNA sequence and no non-specific exonic cut sites were found [18]. Mouse genotyping was performed either: (i) in house via High Resolution Meltcurve (HRM) analysis on the Quant Studio 6 (QS6, Thermo Scientific). Primers used were GGATTCTGAGGCCTTCTGA and CCTCTTACCATGATGTAAGCCA; or (ii) commercially genotyped using real-time PCR (Transnetyx, TN, USA). Further validation of genotypes was performed using an RFLP assay whereby the knock-in removes a TaqI restriction site (primers TGGCTGAAAACCCAAGTACACT and AGCTTGACAATTGTGGGACCA).

Protocols for animal studies were approved by the University of Auckland Animal Ethics Committee and performed in accordance with the NZ Animal Welfare Act (1999) and the Guide for the Care and Use of Laboratory Animals [19] and complied with the ARRIVE guidelines [20]. Animals were maintained in a 12h light/dark cycle with a 30-minute dawn/dusk phase. Temperature was set at 22°C with a tolerance of +/-2°C. Humidity was 55% with a tolerance of +/- 10%. Mice had food and water ad libitum, and the diet was the Teklad 2018, 18% protein rodent diet. Body composition was measured by DXA using a Lunar PIXImus densitometer (DEXA-GE). The machine was calibrated before each session with a phantom provided by the manufacturer. Mice were anesthetized by isoflurane inhalation during the measurement and mouse length was measured in supine position prior to anaesthetic wearing off. Analysis of images was performed with the Lunar PIXImus v2.10 software.

Grip strength was assessed using a Harvard Apparatus Grip Strength Meter with the grid for mice attachment (Harvard Apparatus, Holliston, MA, USA). Each mouse underwent a familiarisation 2 days prior to the experiment and all mice were tested in triplicate taking the average value of the triplicate. Balance/coordination was assessed using the rotarod technique using the RotamexX-5 (Columbus Instruments, USA). Tissues and serum were harvested at euthanasia at around 21.5 months of age. Serum samples were frozen at −80°C until use. Gastrocnemius tissue was preserved in RNALater (Invitrogen) and stored at −20°C prior to mRNA preparation using RNeasy Mini Kit (Qiagen) with pre-treatment with Proteinase K as recommended for fibrous tissue. Quantification of mRNA was done using nanodrop (Thermo Scientific). The presence of genomic-DNA contamination was assessed using power Syber Green RNA to CT one step kit (Thermofisher, 4389986, USA) comparing no-RT enzyme controls to those with RT enzyme present and primers detecting beta-actin (CACTGTCGAGTCGCGTCC and TCATCCATGGCGAACTGGTG). There was no amplification from no-RT control samples. One step TB green primer script RT-PCR kit II (Takara, RR086A, Japan) was used for measuring myostatin by RT-qPCR. Primers used for myostatin were AGAAGATGGGCTGAATCCCT and CATCGCAGTCAAGCCCAAAG. Three reference genes were selected from a test panel of 7; Beta-actin (as above), SDHA (TCGACAGGGGAATGGTTTGG and CTTCCGAGCTTCTGCACCAT), and HPRT (AGTCCCAGCGTCGTGATTAG and TTTCCAAATCCTCGGCATAATGA). Efficiency of all primer pairs were determined using calibration curves and final relative gene expression (RGE) was calculated using the Pfaffle method, with geometric averaging of relative quantities of reference genes [21, 22]. Two-tailed unpaired t-tests were used to assess statistical differences between knockin mice and their wildtype controls.

## RESULTS

To investigate the effect of the A allele of rs373863828 on relative levels of lean and fat mass in humans we performed DXA scans on a cohort of 189 young Māori and Pacific men. Baseline characteristics of the participants by genotype are presented in Table 1. The presence of the rs373863828 A allele had no significant association with bone mineral density, bone mineral content or resting energy expenditure (Table 2). The A allele trended towards association with increase in relative and total lean mass but this was not statistically significant (p=0.06) (Table 2). However, total fat mass was significantly (p=0.031) lower when adjusted for BMI, age and ancestry (Table 2). One possible candidate for such an effect on body composition is myostatin as it is a hormone that opposes muscle development and thus plays a key role in altering the balance between lean and fat mass [23–27]. We found that carriers of rs373863828-A allele had significantly lower plasma concentrations of myostatin when adjusted for BMI, age and ancestry (β −430 pg/ml p=0.04) (Table 2).

**Table 1.**
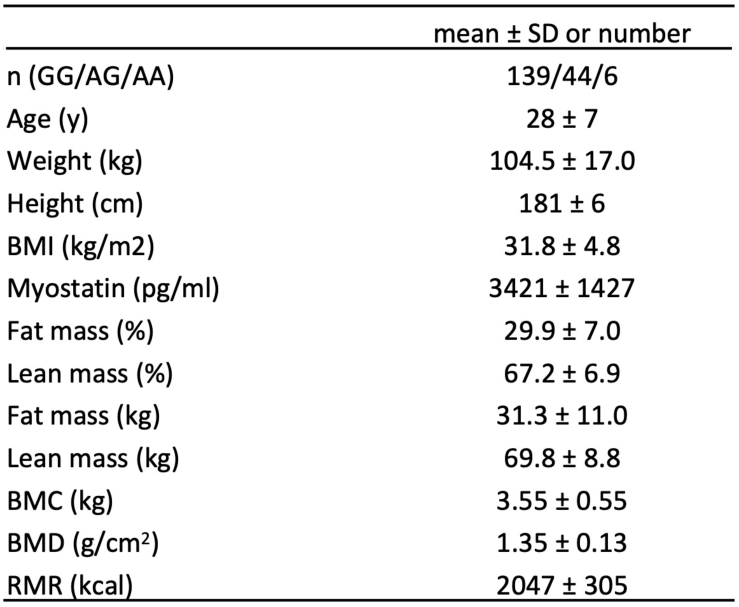
Participant characteristics.

**Table 2.** Impact of rs373863828 A allele on body composition, plasma myostatin, resting metabolic rate (RMR) and bone mineral content and density (BMC and BMD). Association significance was assessed via multivariable linear regression with age, ancestry (ANC) and body mass index (BMI) as covariates. Data are expressed as Beta-coefficients (Beta) ± standard error (SE).

|  | Age, ANC adjusted |  |  | BMI, Age, ANC adjusted |  |  |
| --- | --- | --- | --- | --- | --- | --- |
|  | Beta | SE | p-value | Beta | SE | p-value |
| Myostatin (pg/ml) | -384 | 193 | **0.048** | -430 | 197 | **0.040** |
| Fat mass (%) | -1.13 | 0.96 | 0.240 | -1.19 | 0.74 | 0.062 |
| Lean mass (%) | 1.10 | 0.94 | 0.241 | 1.13 | 0.71 | 0.061 |
| Fat mass (kg) | -1.39 | 1.50 | 0.358 | -1.57 | 0.87 | **0.031** |
| Lean mass (kg) | 1.68 | 1.18 | 0.159 | 1.11 | 0.98 | 0.149 |
| BMC (kg) | 0.09 | 0.07 | 0.228 | 0.08 | 0.07 | 0.252 |
| BMD (g/cm<sup>2</sup>) | 0.02 | 0.02 | 0.218 | 0.02 | 0.02 | 0.240 |
| RMR (kcal) | 38.0 | 43.1 | 0.379 | 32.5 | 38.6 | 0.401 |

Identifying the associations of specific gene variants with body composition in humans is confounded by the diversity of genetic backgrounds, that variants generally have weak effect sizes (reducing study power) and the differences in environmental exposures between different individuals. Therefore, to address the role of rs373863828 on an isogenic background we created two knockin mouse models. CRISPR/Cas9 was used to convert the orthologous mouse CREBRF Arg at position 458 to Gln in both C57 and FVB mice using CRISPR/Cas9 (Suppl. Figure 1). Mating pairs both homozygous for the glutamine allele produced litter sizes that were not different from mating pairs carrying the arginine allele indicating there was no effect on in utero or postnatal survival (data not shown). All mice used in this study came from mating heterozygous mice.

We studied body composition in male mice homozygous for either the arginine or the glutamine allele at 25-weeks (young) and 20-months (old) of age. While the knockin had different effects on overall body weight between the strains a consistent pattern of body composition was observed (Suppl. Table 1). In the younger mice homozygous for the variant there was a trend of higher relative lean body mass on both FVB/NJ and C57Bl/6j backgrounds. However, this was not statistically significant (Figure 1A&B). In the older male C57 knockin mice the pattern of increased relative lean mass was highly significant (p<0.01) (Figure 1C). The glutamine allele also associated with greater grip strength in the 20-month-old male mice (p<0.01) (Figure 1D). We examined levels of myostatin and found that circulating concentrations of this hormone were significantly lower in 20-month old C57 knockin mice (p<0.05) and myostatin mRNA levels in gastrocnemius muscle were also significantly reduced (p<0.05) (Figure 1E-F).

**Figure 1:**
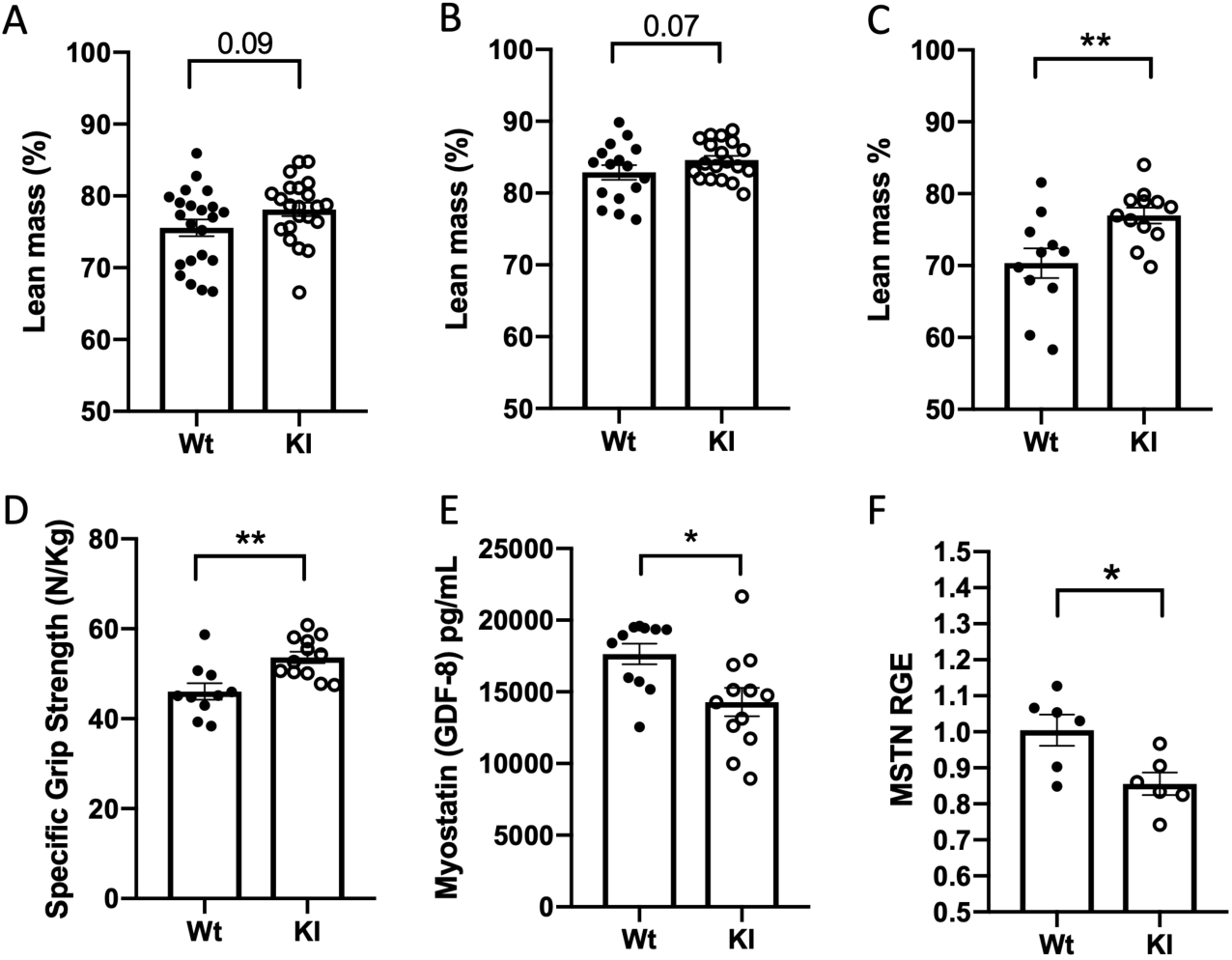
Lean mass expressed as a % of total mass as determined by DXA (Lunar PIXImus densitometer)in C57 male 25-week-old (A) FVB/n male 25-week-old NZ (B) and C57 male 20-month-old (C). Specific grip strength (D) Circulating myostatin (E) gastrocnemius myostatin mRNA relative gene expression (RGE) (F) in 20-month-old C57 male mice.

Since we observed genotype-related differences in body size in the mice (Suppl. Table 1), we also normalised the lean and fat masses to body length (Suppl. Figure 2). These results show that when normalised for body length, lean mass is essentially preserved but fat mass relative to body length is lower.

## DISCUSSION

The minor A allele of rs373863828 SNP in the *CREBRF* gene can be considered a favourable BMI allele in that it is associated with such a large increase in BMI while at the same time associated with a significant decrease in risk of T2D [7, 8]. To provide further insights into the contribution of the CREBRF rs373863828 minor allele to adiposity and lean mass we studied the impact this allele on body composition in a cohort of New Zealand based young healthy men of various Māori and Pacific ancestries. Although the direction of change in relative lean (increase) and fat mass (decrease) was the same as in the recent studies in Samoan studies [15] this was not statistically significant (p=0.06). This pattern would be consistent with previous body composition studies showing that higher relative lean mass for a given BMI is observed in Māori and Pacific peoples compared to people of European or Asian ancestry in New Zealand [12, 13]. The most plausible reason for our lack of statistical significance in our studies is inadequate power. It is also possible that age may influence the effect of rs373863828 has on body composition since the more significant effects reported previously in Samoan subjects were in an older cohort (48-53 years) [15] compared with the cohort reported here (average age of 28 years).

A strength of the current study is that we are able to analyse body composition in knockin mouse models and at two different ages, as well as studying the impact of the variant in two different isogenic backgrounds. Further, it allowed us to study the impact in the context of being homozygous for the variant and without confounding influences of environment. Notably the effects seen are more obvious in mice whose age was equivalent to late middle age in humans [28] which is consistent with the observations in humans [15]. These mouse studies provide independent evidence to support the case that the p.Arg457Gln coding variant of CREBRF promotes an increase in relative lean mass and is consistent with the observation that the rs373863828 minor allele is associated with a reduced risk of T2D. This is because muscle is the major site of insulin-stimulated glucose disposal so small changes in mass can have an effect on glucose disposal [29] and large-scale studies have shown that increases in muscle mass associate with improvements in glucose metabolism and reduced risk of T2D [30]. Given that CREBRF and its target CREB3 are widely expressed other mechanisms are likely to also contribute to the reduced risk of T2D. For example, we recently demonstrated that the rs373863828 A allele associates with increased plasma insulin in response to a glucose load in males, independent of differences in insulin sensitivity [16].

An important finding of the current study is the association of the rs373863828 A allele with reduction in myostatin levels in both the mice and in the human participants in our study. This is likely to contribute to the increase in relative lean mass as lower myostatin is associated with increased in muscle mass [23, 31] and reductions in fat [26]. It is known that lower levels of myostatin result in increases in relative lean mass with ageing which is consistent with our findings [32]. In addition, reduced myostatin levels are also associated with a lower risk of T2D [23, 24, 33] which is consistent with the effects of the rs373863828 A allele on risk of T2D. Moreover, reduced myostatin expression has been associated with increased muscle mass in other mammals [34]

To date two main roles have been identified for CREBRF; the first is to bind to the transcription factor CREB3 and regulate CREB3 levels and activity [6]. The transcription factor component CREB3 is cleaved and thus activated by ER/Golgi stress [35] and in turn plays a key role in regulating endoplasmic and Golgi stress responses in cells [36, 37]. CREBRF has also been shown to regulate the cellular location and activity of the glucocorticoid receptor [38]. The rs373863828 A allele could affect myostatin, either through glucocorticoid- or ER/Golgi stress-mediated pathways. Both ER stress [39] and glucocorticoid signalling [40] are known to play important roles in regulating muscle mass. ER stress induces expression of myostatin [41] and it is notable that the myostatin promoter contains a CREB responsive element [42] thus potentially allowing it to be regulated by CREB3. ER stress also impairs myostatin processing thus resulting in reduced release of the mature form [43]. Changes in myostatin levels also contribute to reduced muscle mass downstream of glucocorticoids and this contributes to the muscle wasting effects of corticosteroids [40, 44]. The myostatin gene promoter contains a glucocorticoid response element that allows the gene to be upregulated by glucocorticoids [44] and in mice myostatin levels are elevated *in vivo* by glucocorticoid treatment [40]. While glucocorticoids have a wide range of effects, myostatin appears to be crucial for the effects on muscle as myostatin knockout mice are resistant to glucocorticoid-induced muscle atrophy [45]. Collectively this suggests plausible links between CREBRF and regulation of myostatin levels.

One limitation of our study is that our human cohort did not include females. Given that it is known there is sexual dimorphism in the regulation of myostatin [25, 31, 46] it is possible that different effects will be observed in women. Other sexually dimorphic effects of rs373863828 have been observed, for example the male-specific effects observed on height [17, 47] and differences in the pattern of body composition between males and females [15]. Further studies will be required to resolve these issues. Our results also provide further insights into the thrifty gene hypothesis [48, 49]. One interpretation of this hypothesis is that there is positive selection for fat accumulation as an energy store for long periods of starvation [50]. In the case of rs373863828 evidence for natural selection occurring around this locus in Polynesian people has been suggested as evidence that this is a thrifty gene [8]. Our results do not support this as there is no evidence it is driving an increase in adiposity. Similarly, previously a variant of PPARGC1 was proposed as a thrifty gene in Polynesian people [51] but this has been rejected following further analysis of the locus in the genomes of Polynesian people for a signature of selection [52]. Moreover, this hypothesis has previously been questioned on the basis of logic and lack of evidence [53]. Reasons for natural selection at the CREBRF locus will require further study and other hypotheses can be proposed. For example, it has been suggested that recent genetic selection in indigenous populations may have been caused by the arrival of novel Western diseases [54]. Given the known roles of CREB3 and CREBRF in regulating viral infectivity [55, 56] it is plausible that recent natural selection at the CREBRF locus relates to such effects.

## CONCLUSIONS

Using mouse models and human phenotyping we provide evidence that in males the CREBRF p.457Gln allele does not drive increased adiposity and is hence not likely to be a thrifty gene as previously proposed [8]. Our results are consistent with a model where increases in relative lean mass would make the greatest contribution to the increases in mass that cause increases in BMI, and this is supported by the observation of lower myostatin levels in participants with a p.457Gln allele and knockin mice that model this variant. Such a phenotype is consistent with improvements in glucose metabolism and consequent reduction in risk of T2D in carriers of the CREBRF p.457Gln allele despite their increased BMI [24, 33, 57].

## Data Availability

Data is available on reasonable request

## ACKNOWLEDGEMENTS

He mihi nui tēnei ki ngā kai tuku taonga (we would like to thank the study participants for their time and tissue samples).

## Credit authorship contribution statement

**Kate Lee** Conceptualization, Supervision, Formal analysis, Writing – original draft, review & editing, Investigation **Sanaz Vakili** Methods, Investigation.**Hannah Burden** Methods, Investigation **Shannon Adams** Methods, Investigation **Greg C. Smith** Methods **Braydon Kulatea** Investigation **Robert D. Atiola** Methods, Investigation **Conor Watene-O’SuIllivan** Investigation, conceptualization **Morag Wright-McNaughton** Investigation **Danielle Sword** Investigation, **Ryan Paul** Writing - review & editing **Phillip Wilcox** Writing - review & editing, **Prasanna Kallingappa** Methods **Jeremy D. Krebs** Conceptualization, Writing - review & editing. **Rosemary M. Hall** Conceptualization, Writing - review & editing. **Lindsay D. Plank** Conceptualization, Writing - review & editing. **Tony R. Merriman** Conceptualization, Writing - review & editing. **Rinki Murphy** Conceptualization, Writing - review & editing. **Troy L. Merry** Conceptualization, Supervision, Formal analysis, Writing – original draft, review & editing, Investigation **Peter R. Shepherd** Supervision, Formal analysis, conceptualization, Writing – original draft, review & editing.

---

**Supplementary Table 1.** Body composition characteristics of CREBRF KI vs WT mice. FVB/n and C57 male mice at 25 weeks of age and C57 male mice at 20 months of age. TM: total mass (g), L-NA: length nose-anus (mm), LM: lean mass (g), FM: fat mass (g), L%: LM as a % of TM, F%: FM and a % of TM. Statistics were two-way unpaired t-test.

|  |  |
| --- | --- |
| TM | total mass, g |
| L-NA | Length, cm (nose to anus) |
| LM | lean mass, g |
| FM | fat mass, g |
| L% | LM as % of TM |
| F% | FM as % of TM |

|  |  | Wt |  | KI |  | Difference | P-value |
| --- | --- | --- | --- | --- | --- | --- | --- |
|  |  | n | Mean (SD) | n | Mean (SD) |  |  |
| C57BL/6j<br>25-week | TM | 21 | 32.0 (2.7) | 21 | 30.8 (3.3) | -1.18 ± 0.93 | 0.062 |
|  | L-NA | 21 | 95.5 (2.3) | 21 | 94.8 (2.7) | 0.67 ± 0.78 | 0.4 |
|  | LM | 21 | 22.4 (1.7) | 21 | 22.8 (1.6) | 0.50 ± 0.51 | 0.325 |
|  | FM | 21 | 7.4 (2.1) | 21 | 6.1 (1.4) | -1.29 ± 0.56 | **0.0255** |
|  | L% | 21 | 75.6 (5.4) | 21 | 78.1 (4.3) | 2.57 ± 1.48 | 0.09 |
|  | F% | 21 | 24.3 (5.5) | 21 | 22.1 (4.4) | -2.19 ± 1.50 | 0.15 |
| FVB/n<br>25-week | TM | 16 | 30.9 (2.5) | 19 | 30.2 (2.2) | -0.75 ± 0.79 | 0.17 |
|  | L-NA | 16 | 98.4 (2.8) | 19 | 99.2 (3.0) | -0.78 ± 0.98 | 0.4 |
|  | LM | 16 | 23.9 (1.4) | 19 | 23.9 (1.8) | 0.03 ± 0.56 | 0.95 |
|  | FM | 16 | 5.0 (1.5) | 19 | 4.4 (1.0) | -0.64 ± 0.43 | 0.14 |
|  | L% | 16 | 82.9 (4.0) | 19 | 84.6 (2.6) | 1.70 ± 1.14 | 0.072 |
|  | F% | 16 | 17.4 (3.8) | 19 | 15.4 (2.6) | -1.96 ± 1.08 | 0.079 |
| C57BL/6j<br>20-month | TM | 11 | 38.5 (5.3) | 12 | 32.6 (2.4) | -5.9 ± 1.7 | **0.0022** |
|  | L-NA | 11 | 101.7 (3.1) | 12 | 97.4 (3.0) | -4.3 ± 1.28 | **0.0029** |
|  | LM | 11 | 25.8 (2.6) | 12 | 23.8 (1.0) | -1.92 ± 0.80 | **0.025** |
|  | FM | 11 | 11.3 (4.2) | 12 | 7.2 (1.7) | -4.03 ± 1.30 | **0.0054** |
|  | L% | 11 | 70.3 (6.9) | 12 | 77 (3.8) | 6.63 ± 2.28 | **0.0085** |
|  | F% | 11 | 29.7 (6.8) | 12 | 23.5 (3.9) | -6.13 ± 2.30 | **0.0143** |

**Supplementary Figure 1.**
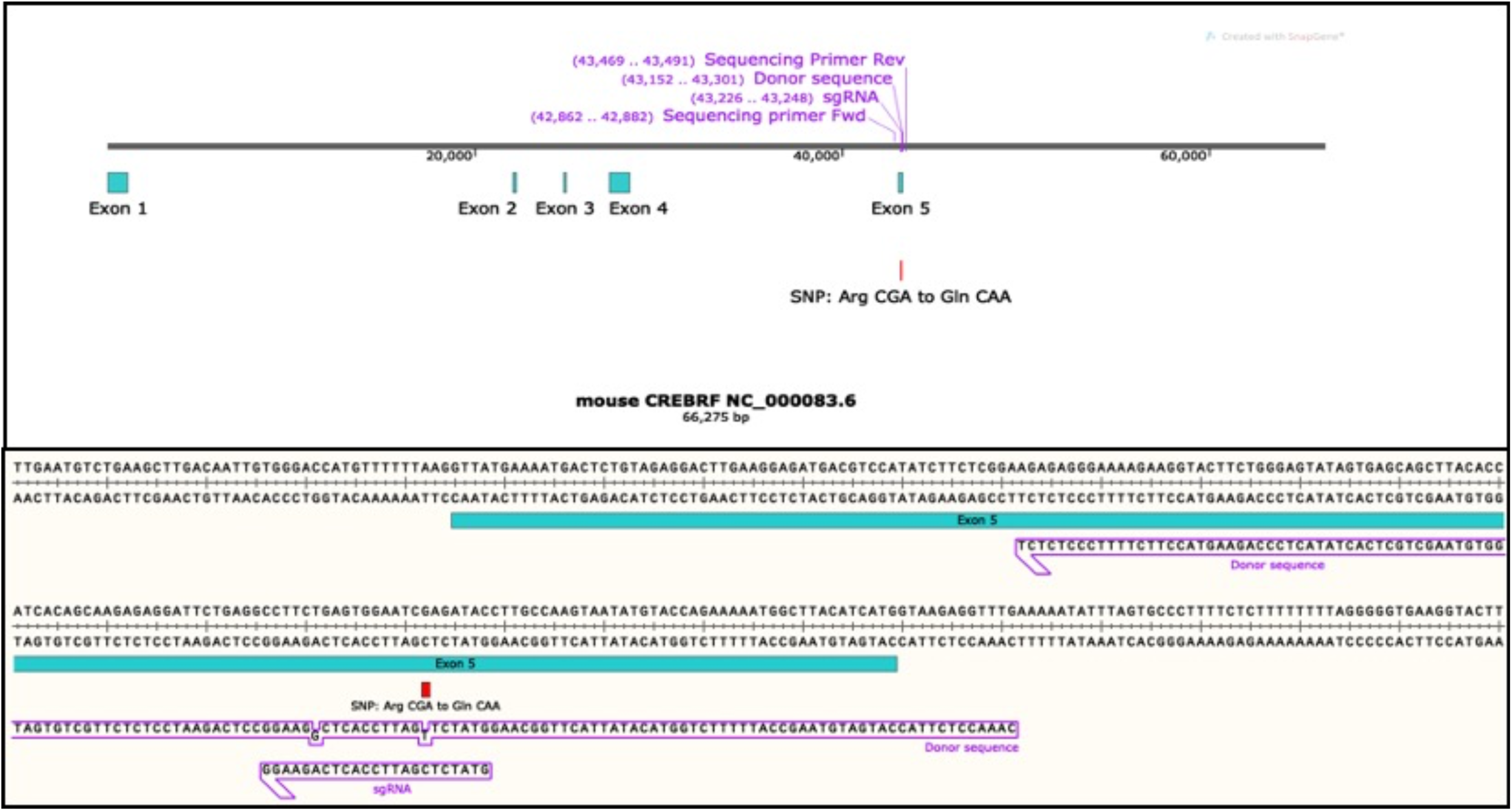
Mouse CREBRF gene sequence showing CrispR Cas9 gRNA and HDR donor sequence and position of SNP. Image created with Snapgene ®.

**Suppl Figure 2:**
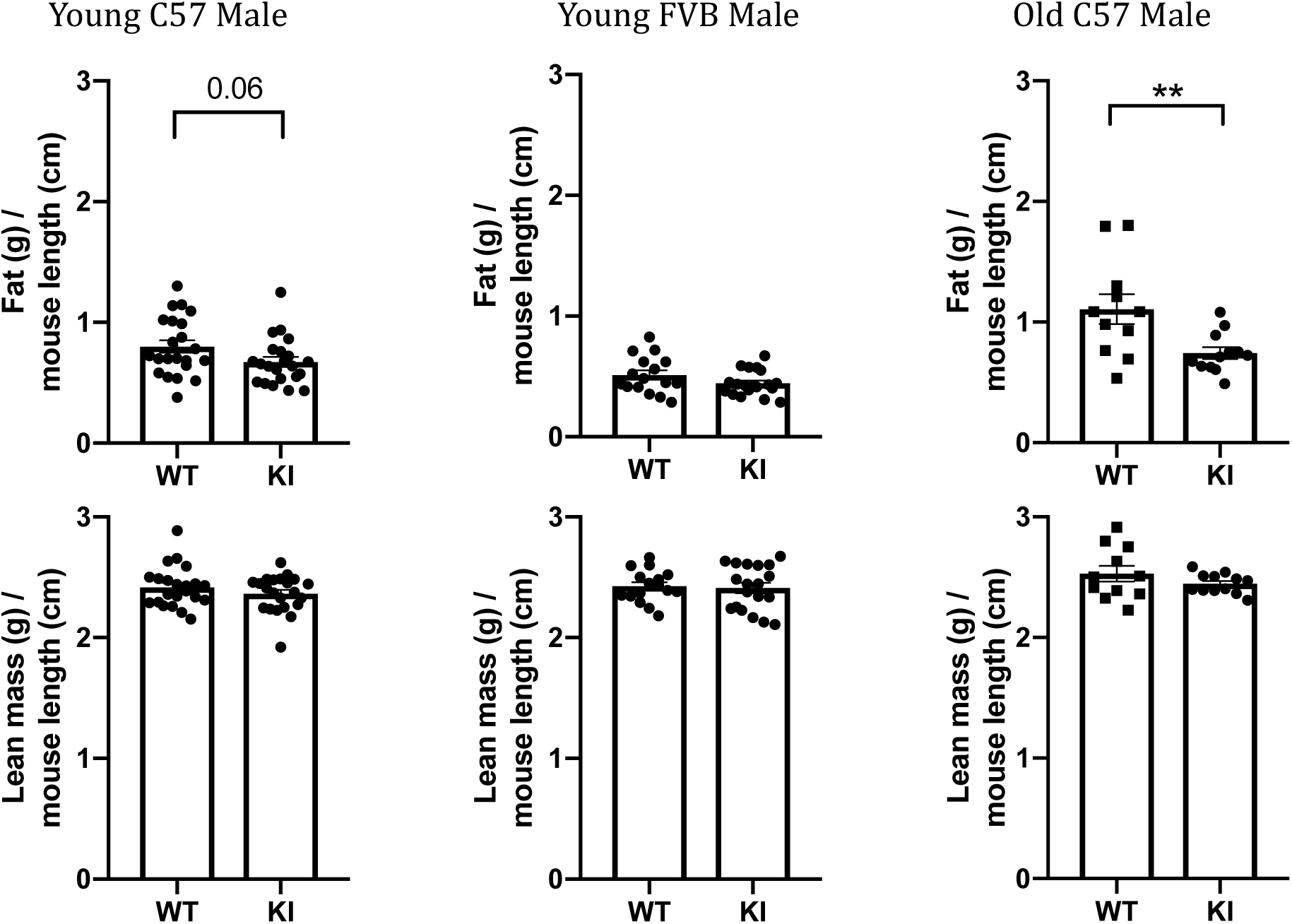
Lean mass (g) and fat mass (g) corrected for mouse length (cm) in FVB/n and C57 male mice at 25 weeks of age and C57 male mice at 20 months of age. Statistics were two-way unpaired t-test.

